# Prospective Multicenter Evaluation of the QuickNavi-Campylobacter Assay in Stool Specimens

**DOI:** 10.64898/2026.03.03.26346362

**Authors:** Shinji Hatakeyama, Yumi Hirose, Yusaku Akashi, Tomoka Kusama, Naoto Ishimaru, Eri Morimoto, Satoru Iwashima, Kenji Suzuki, Keisuke Enomoto, Satoshi Suzuki, Michiko Sekine, Tatsuo Nishimura, Norihiko Terada, Miho Takahashi-Igari, Mariko Abe, Kota Yamada, Daisuke Kato, Kiyofumi Ohkusu, Hiromichi Suzuki

## Abstract

The rapid diagnosis of *Campylobacte*r infections is important for the management of infectious gastroenteritis. Although stool culture is considered the gold standard, its sensitivity is limited and it requires prolonged incubation times. We performed a prospective multicenter study at nine healthcare facilities in Japan to evaluate a *Campylobacter* rapid antigen test using stool specimens between March 2024 and August 2025. Patients with suspected infectious gastroenteritis were consecutively enrolled and tested using QuickNavi-Campylobacter and compared with the FilmArray Gastrointestinal Panel as the reference method. Discordant results were further evaluated by culturing and additional PCR assays. In total, 410 patients were included in the final analysis. The positive, negative, and total concordance rates between QuickNavi-Campylobacter and FilmArray Gastrointestinal Panel were 79%, 99%, and 93%, respectively. The positive concordance rate decreased in specimens collected ≥ 6 days after the onset of symptoms (50%). QuickNavi-Campylobacter demonstrated relatively good concordance with the FilmArray Gastrointestinal Panel in a real-world multicenter setting. These results suggest that this rapid antigen test may be particularly useful for the early diagnosis of suspected campylobacteriosis.

## 1. Introduction

*Campylobacter* species are Gram-negative, curved, or spiral-shaped motile bacteria that grow under microaerophilic conditions and commonly colonize the gastrointestinal tract of a wide range of animals [1]. Among these species, *Campylobacter jejuni* and *Campylobacter coli* are the predominant pathogens causing human campylobacteriosis [2]. In industrialized countries, *Campylobacter* species are recognized as the leading bacterial cause of acute gastroenteritis, exceeding the incidence of other bacterial pathogens, such as *Salmonella* and *Shigella* [3].

Traditionally, stool culture has been regarded as the gold standard for the diagnosis of campylobacteriosis. However, *Campylobacter* culture requires specialized culture media and incubation under specific conditions, which limits its clinical utility owing to the time required to obtain results [4, 5]. In addition, testing methods differ among laboratories, the diagnostic accuracy of the stool culture varies accordingly [5, 6]. In recent years, rapid molecular diagnostic tests, including multiplex gastrointestinal panels, have been introduced for the diagnosis of infectious gastroenteritis, and have demonstrated higher sensitivity for *Campylobacter* detection in comparison to conventional culture methods [7–9]. Nevertheless, these molecular assays are costly and require specialized equipment, limiting their availability to selected medical facilities [9].

More recently, immunochromatographic assays enabling the direct detection of *Campylobacter* in stool specimens have become clinically available.

QuickNavi™-Campylobacter (Denka Co., Ltd., Tokyo, Japan) is a newly developed rapid antigen test that can be performed using a workflow similar to that of influenza diagnostic kits, providing results within approximately 15 min. In our previous study comparing QuickNavi-Campylobacter with conventional culture methods, positive and negative concordance rates were 93% and 99%, respectively [10]. However, that study was limited by its single-center design and use of conventional stool culture as the primary reference method. Therefore, further multicenter evaluations using a highly sensitive molecular reference method, such as the FilmArray Gastrointestinal Panel (FilmArray GI Panel; bioMérieux, Marcy-l’Étoile, France), are warranted to establish the diagnostic accuracy of rapid antigen tests for campylobacteriosis.

Accordingly, we performed a multicenter evaluation to assess the diagnostic performance of QuickNavi-Campylobacter in comparison to the FilmArray GI Panel.

## 2. Patients and methods

This prospective multicenter study was performed at nine healthcare facilities in Japan, comprising five acute care hospitals (University of Tsukuba Hospital, Akashi Medical Center, Chutoen General Medical Center, Tone Chuo Hospital and Tsukuba Medical Center Hospital) and four clinics (Akashi Internal Medicine Clinic, Enomoto Children’s Clinic, Nishimura Pediatric Clinic, Tsuchiura Beryl Clinic). Patients suspected of having infectious gastroenteritis between March 2024 and August 2025 were enrolled consecutively. Written informed consent was obtained from all participants or their legally authorized representatives. The study protocol was approved by the Ethical Committee of the University of Tsukuba Hospital (approval number: R05-028).

### 2.1. Study process

During the study period, patients who visited participating healthcare facilities and were clinically suspected of having infectious gastroenteritis by physicians were enrolled. After obtaining written informed consent, stool specimens were collected at each facility. Clinical signs and symptoms were recorded, and a *Campylobacter* rapid antigen test using QuickNavi-Campylobacter was performed on stool specimens, in accordance with the manufacturer’s instructions. Healthcare professionals interpreted the test results.

As a reference method, the FilmArray GI Panel was promptly performed at facilities where testing was available. At facilities where immediate testing was not feasible, stool specimens were stored at −30°C or lower and transported under frozen conditions to University of Tsukuba Hospital, where FilmArray GI Panel testing was subsequently conducted.

For specimens with discordant results between the antigen test and FilmArray GI Panel, additional analyses were performed using residual stool specimens.

*Campylobacter* culture was conducted using AccuRate™ Skirrow Agar Modified (Shimadzu Diagnostics Corporation, Tokyo, Japan) under microaerophilic conditions at 35°C for 72 h. When bacterial growth was observed, species identification was performed using a VITEK^®^ MS PRIME (bioMérieux, Marcy-l’Étoile, France). For culture-negative specimens, Species-specific PCR assays targeting *Campylobacter jejuni*, *Campylobacter coli*, and *Campylobacter upsaliensis* were performed according to previously reported methods [11].

### 2.2. Statistical analyses

The positive, negative, and total concordance rates, as well as the positive and negative predictive values of the antigen tests were calculated with 95% confidence intervals (CIs) using the Clopper-Pearson method. The degree of agreement between the two tests was evaluated by calculating the Cohen’s kappa coefficient. All statistical analyses were performed using IBM SPSS Statistics (ver. 29.0.2.0, IBM, Armonk, NY, USA).

## 3. Result**s**

In this multicenter study, 422 patients provided their written informed consent for participation. Six participants were excluded because they were unable to obtain stool specimens, and five were excluded because they were unable to perform the reference method (FilmArray GI Panel). In addition, one participant was excluded after a review of clinical information revealed that the individual was asymptomatic. Consequently, 410 participants were included in this analysis.

The clinical characteristics of the participants are summarized in Table 1. The median age was 23 years (range, 0–88 years), and 40% of the patients were female. The participants were categorized by age as follows: ≤ 6 years (98, 24%), 7–17 years (39, 10%), and ≥ 18 years (273, 67%). The median interval from symptom onset to specimen collection was one day. Diarrhea was the most frequent symptom (97%), followed by abdominal pain (66%), and fever (41%).

**Table 1.** Patient characteristics (n=410)

|  | Number (%) |
| --- | --- |
| Age (median [range]) | 23 [0-88] |
| Female | 162 (39.5) |
| Prior antimicrobial use | 29 (7.1) |
| Recent history of international travel (<1 month) | 40 (9.8) |
| Symptomatic household contacts | 71 (17.3) |
| Days from symptom onset to specimen collection (median [IQR]) | 1 [1-3] |
| Signs and symptoms |  |
| Fever ($\geq 37.8^{\circ}\text{C}$) | 166 (40.5) |
| Chills | 101 (24.6) |
| Diarrhea | 398 (97.1) |
| Abdominal pain | 269 (65.6) |
| Vomiting | 138 (33.7) |
| Hematochezia | 28 (6.8) |

IQR, interquartile range

Table 2 summarizes the comparison between the QuickNavi-Campylobacter antigen test and FilmArray GI Panel using stool specimens. The positive, negative, and total concordance rates were 79, 99, and 93%, respectively.

**Table 2.** Comparison between QuickNavi-Campylobacter and FilmArray.

|  |  | FilmArray |  |
| --- | --- | --- | --- |
|  |  | Gastrointestinal Panel |  |
|  |  | Positive | Negative |
| *Campylobacter* rapid antigen test | Positive | 89 | 3 |
| (QuickNavi-Campylobacter) | Negative | 24 | 294 |
| Positive concordance rate (%) |  | 78.8 (70.1-85.9) |  |
| Negative concordance rate (%) |  | 99.0 (97.1-99.8) |  |
| Total concordance rate (%) |  | 93.4 (90.6-95.6) |  |
| Positive predictive value (%) |  | 96.7 (90.8-99.3) |  |
| Negative predictive value (%) |  | 92.5 (89.0-95.1) |  |

Cohen's Kappa coefficient between the two tests was 0.83
The positive, negative, and total concordance rate, positive and negative predictive value are presented with their 95% confidence intervals.

The diagnostic performance of the QuickNavi-Campylobacter test stratified by the number of days from the onset of symptoms to specimen collection is shown in Figure 1. The cumulative positive concordance rate for specimens collected within 1 day of symptom onset was 83% (40/48). In contrast, the positive concordance rate for specimens collected ≥ 6 days after symptom onset was 50% (3/6).

**Figure 1.**
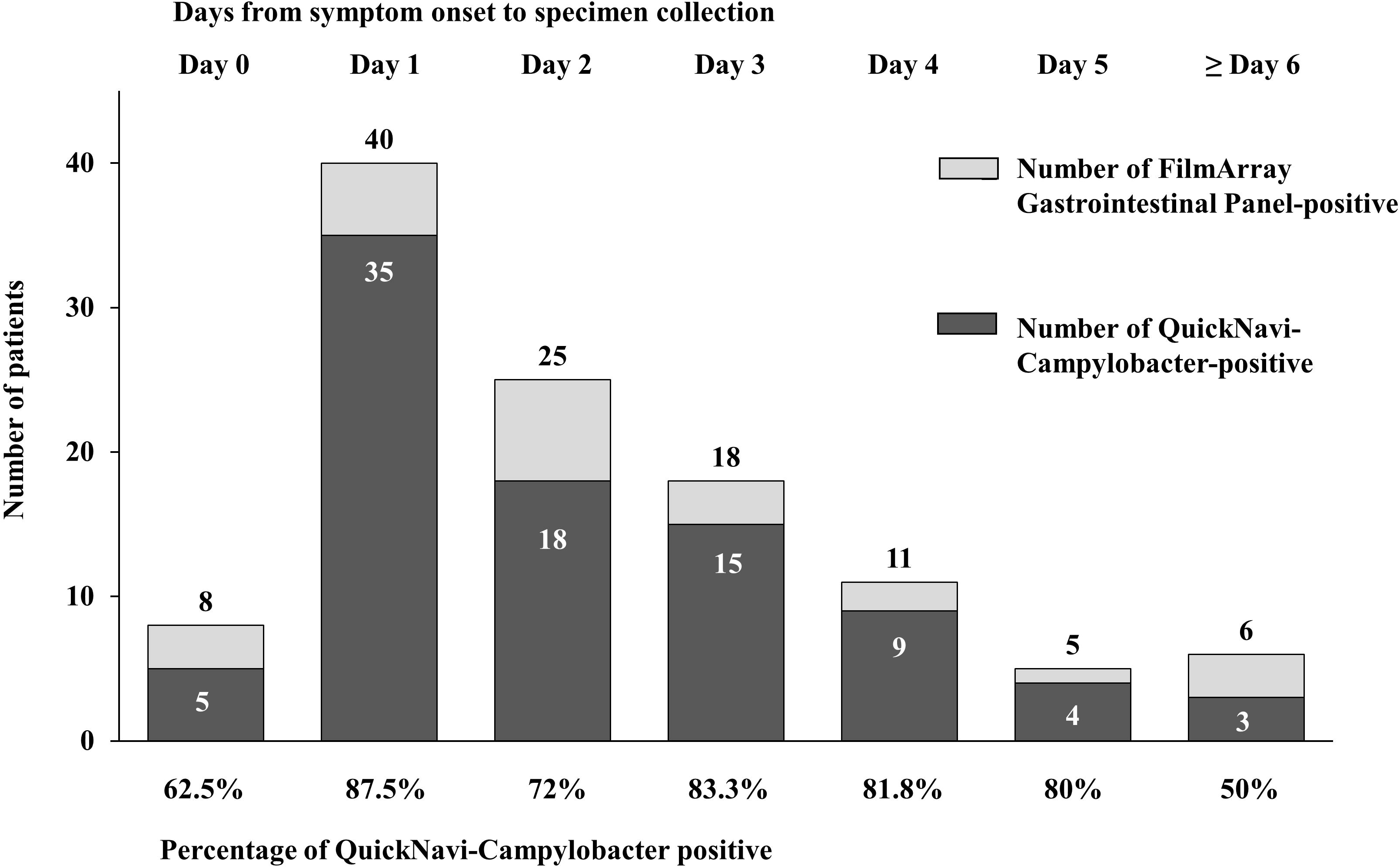
The diagnostic performance of the QuickNavi-Campylobacter test stratified by days from symptom onset to specimen collection.

The 27 cases with discordant results between the antigen test and FilmArray GI Panel are listed in Table 3. Three specimens were positive by the QuickNavi-Campylobacter antigen test, but negative by the FilmArray GI Panel; all three specimens were negative by additional culture and PCR testing. Conversely, 24 specimens were negative by the QuickNavi-Campylobacter antigen test but positive by the FilmArray GI Panel. Among these, *Campylobacter jejuni* and *Campylobacter coli* were identified in eight specimens by culturing and in five specimens by an additional PCR assay. The detailed results for the 410 participants are presented in Supplementary Table 1.

**Table 3.**
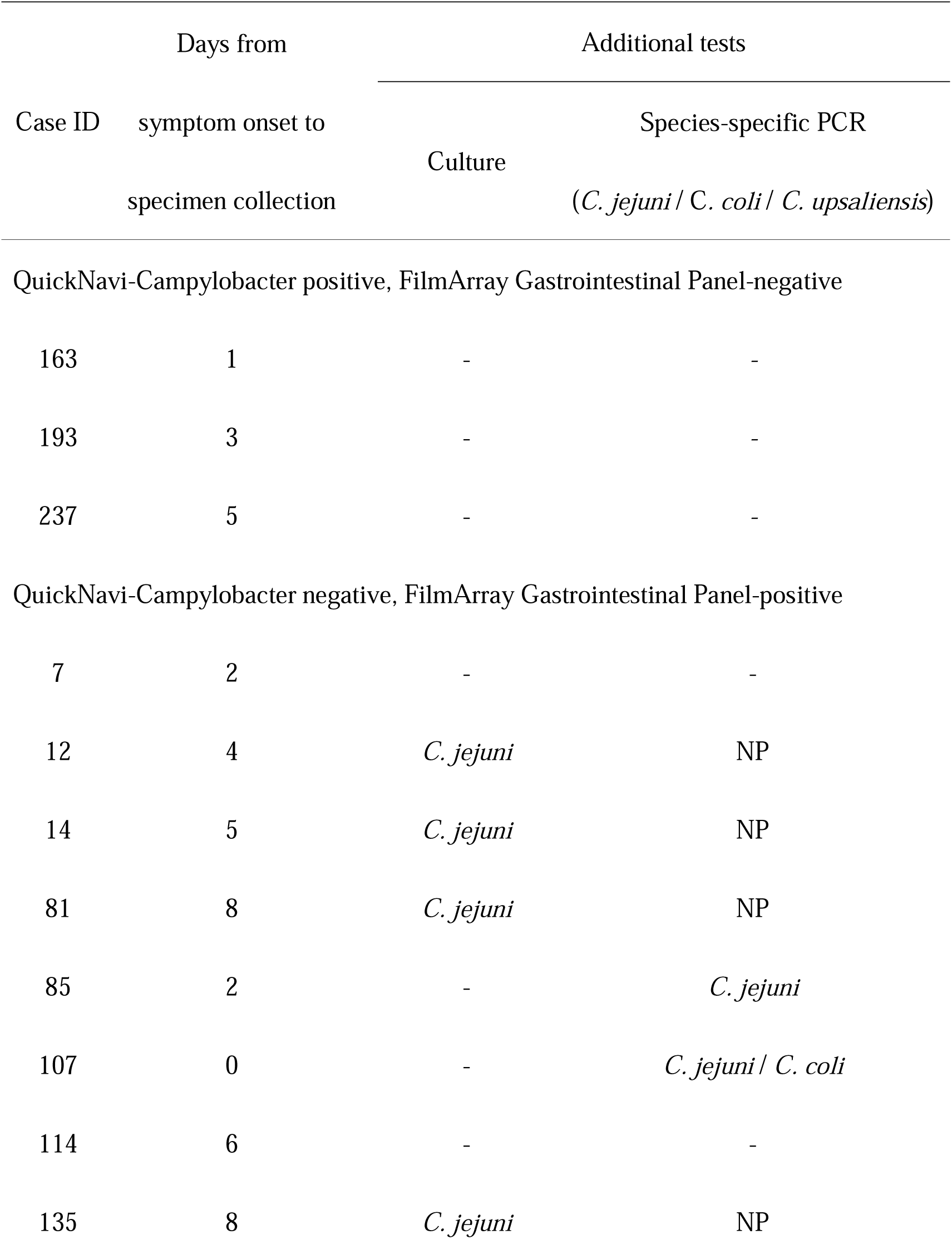
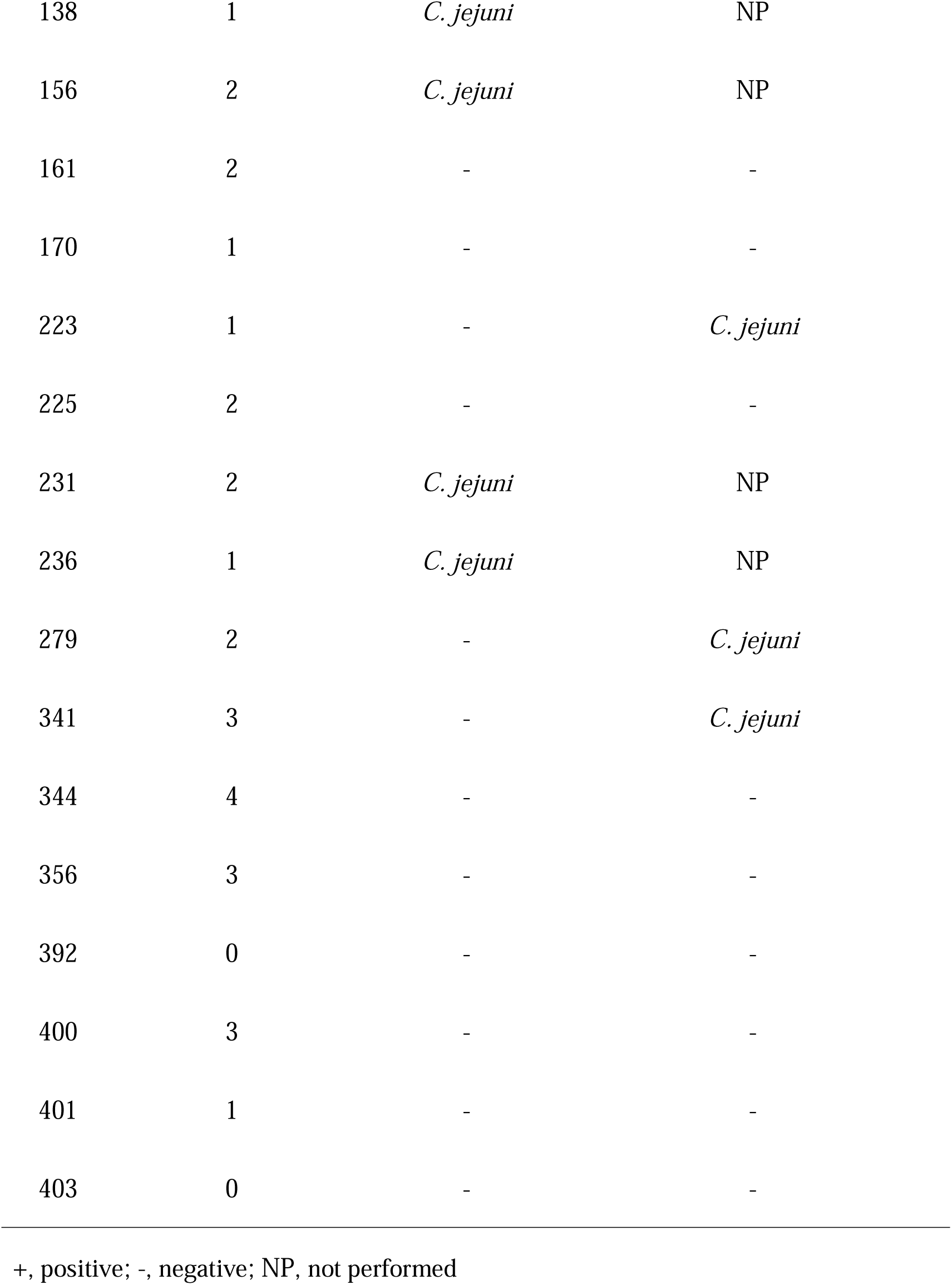
Discrepancy analysis between QuickNavi-Campylobacter and FilmArray.

## Discussion

In this prospective multicenter study, we examined the performance of QuickNavi-Campylobacter across acute care hospitals and clinics, including both pediatric and adult populations. Among the 410 cases included in the analysis,

QuickNavi-Campylobacter showed a positive concordance rate of approximately 80% relative to the FilmArray GI Panel. The positive concordance rate appeared to be lower in specimens collected ≥ 6 days after symptom onset.

*Campylobacter* spp. have been reported to remain detectable in stool for a prolonged period by nucleic acid amplification tests in cases of campylobacteriosis [12]. However, the bacterial load is generally low, and the culture-positive rate of *Campylobacter* spp. in stool samples decreases progressively over time [13] with approximately one-third of patients becoming stool culture–negative within one week after symptom onset [14, 15].

Consistent with these observations, our analysis demonstrated that the current antigen-based assay exhibited sufficient diagnostic performance when applied to stool specimens collected within six days after the onset of disease.

In a previous single-center evaluation of the QuickNavi-Campylobacter assay, the sensitivity was 93% when stool culture was used as the reference method [10]. The FilmArray assay has demonstrated a higher sensitivity for the detection of

*Campylobacter* spp. than stool culture and other molecular assays [9, 16–18]. Previous studies have reported that the sensitivity of stool culture ranges from 60% to 86% relative to the FilmArray assay [7–9, 19], supporting its use as a highly sensitive reference method. In a recent study, false-positive results associated with the FilmArray assay were reported [20]. In our analysis, 11 discordant specimens were negative on both culturing and additional PCR testing. Of these, three exhibited melting curve patterns identical to those previously reported to be associated with false-positive results (Supplementary Figure 1), suggesting that the true diagnostic sensitivity of the antigen assay may differ from that estimated when FilmArray was used as the sole reference standard.

In addition to QuickNavi-Campylobacter, several other rapid assays for the direct detection of *Campylobacter* spp. in stool specimens are commercially available worldwide, including ImmunoCard STAT!^®^ CAMPY (Meridian Bioscience Inc., Cincinnati, OH, USA), and RIDA^®^QUICK Campylobacter (R-Biopharm AG, Darmstadt, Germany). Reported sensitivities for these assays generally range from approximately 70% to 90%, while specificities often exceed 95% [21–23]. However, these findings should be interpreted with caution because of differences in reference methods, patient populations, and specimen collection timing among studies. Thus, direct comparative analyses are warranted for the analytical evaluation of

*Campylobacter* antigen testing.

This study has several limitations. First, antigen test results were interpreted at each participating facility and a double-checking procedure was not strictly defined. Because visually interpreted antigen test results may vary among technicians [24], the current findings might have been influenced by observer bias. In addition, the detection performance of *Campylobacter* spp. other than *Campylobacter jejuni* and *Campylobacter coli* has not yet been fully evaluated. Further investigations using well-characterized isolates, including the less common *Campylobacter* species, are warranted to comprehensively assess the analytical performance of the assay.

In conclusion, the QuickNavi-Campylobacter antigen test demonstrated relatively good concordance with the FilmArray GI Panel in this prospective, multicenter study. These findings suggest that the diagnostic performance of the assay may be particularly favorable in the early phase of illness and may support timely clinical decision-making without the need for advanced infrastructure.

## Supporting information

upplementary Table 1 and Supplementary Figure 1

## Data Availability

All data produced in the present study are available upon reasonable request to the authors

## Acknowledgments

We thank all participating medical institutions for providing clinical information from their patients and for their support with laboratory testing.

## Authorship Statement

All authors meet the ICMJE authorship criteria.

Shinji Hatakeyama collected the samples, conducted laboratory evaluations, performed the statistical analysis, and drafted the manuscript.

Yumi Hirose, Yusaku Akashi, Naoto Ishimaru, Satoru Iwashima, Keisuke Enomoto, Satoshi Suzuki, Tatsuo Nishimura, Norihiko Terada, Miho Takahashi-Igari, and Kota Yamada collected samples and revised the manuscript.

Tomoka Kusama, Eri Morimoto, Kenji Suzuki, Michiko Sekine, and Mariko Abe collected the samples, conducted laboratory evaluations, and revised the manuscript. Daisuke Kato interpreted the results.

Kiyofumi Ohkusu conducted laboratory evaluations and revised the manuscript. Hiromichi Suzuki drafted the manuscript, designed the study, and supervised the project.

