## Supplementary material for "Prospective Multicenter Evaluation of the QuickNavi-Campylobacter Assay in Stool Specimens": upplementary Table 1 and Supplementary Figure 1

| Supplement Table 1. Detailed results of the 410 participants | | | | | | | | | |
| --- | --- | --- | --- | --- | --- | --- | --- | --- | --- |
| Case ID | Age | Sex | Days from symptom onset to specimen collection |  | Test results | |  | Discrepancy analysis | |
|  |  |  |  |  | FilmArray Gastrointestinal Panel | QuickNavi-Campylobacter |  | Culture | *C. jejuni* / *C. coli*/ *C. upsaliensis*  Specific PCR |
| 1 | ≥ 18 | M | 2 |  | - | - |  | NP | NP |
| 2 | ≥ 18 | M | 9 |  | + | + |  | NP | NP |
| 3 | ≥ 18 | M | 0 |  | - | - |  | NP | NP |
| 4 | ≥ 18 | M | 25 |  | - | - |  | NP | NP |
| 5 | ≥ 18 | F | 4 |  | - | - |  | NP | NP |
| 6 | ≤ 6 | F | 2 |  | - | - |  | NP | NP |
| 7 | ≥ 18 | M | 2 |  | + | - |  | *-* | - |
| 8 | ≥ 18 | M | 2 |  | - | - |  | NP | NP |
| 9 | ≥ 18 | M | 4 |  | - | - |  | NP | NP |
| 10 | ≥ 18 | F | 7 |  | - | - |  | NP | NP |
| 11 | ≥ 18 | F | 5 |  | - | - |  | NP | NP |
| 12 | ≥ 18 | F | 4 |  | + | - |  | *C. jejuni* | NP |
| 13 | ≥ 18 | F | 2 |  | - | - |  | NP | NP |
| 14 | ≥ 18 | F | 5 |  | + | - |  | *C. jejuni* | NP |
| 15 | ≥ 18 | F | 3 |  | + | + |  | NP | NP |
| 16 | ≥ 18 | F | 4 |  | + | + |  | NP | NP |
| 17 | ≥ 18 | F | 2 |  | - | - |  | NP | NP |
| 18 | ≥ 18 | F | 3 |  | + | + |  | NP | NP |
| 19 | ≥ 18 | M | 3 |  | - | - |  | NP | NP |
| 20 | ≥ 18 | M | 1 |  | - | - |  | NP | NP |
| 21 | ≥ 18 | M | 6 |  | - | - |  | NP | NP |
| 22 | ≥ 18 | M | 4 |  | + | + |  | NP | NP |
| 23 | ≥ 18 | M | 3 |  | - | - |  | NP | NP |
| 24 | ≥ 18 | F | 2 |  | - | - |  | NP | NP |
| 25 | ≥ 18 | F | 10 |  | - | - |  | NP | NP |
| 26 | ≥ 18 | F | 2 |  | - | - |  | NP | NP |
| 27 | ≥ 18 | M | 78 |  | - | - |  | NP | NP |
| 28 | ≥ 18 | M | 6 |  | - | - |  | NP | NP |
| 29 | ≥ 18 | F | 2 |  | - | - |  | NP | NP |
| 30 | ≥ 18 | M | 1 |  | - | - |  | NP | NP |
| 31 | ≥ 18 | F | 3 |  | - | - |  | NP | NP |
| 32 | ≥ 18 | F | 2 |  | - | - |  | NP | NP |
| 33 | ≥ 18 | F | 0 |  | - | - |  | NP | NP |
| 34 | ≤ 6 | F | 7 |  | - | - |  | NP | NP |
| 35 | ≤ 6 | M | 2 |  | - | - |  | NP | NP |
| 36 | ≥ 18 | F | 1 |  | - | - |  | NP | NP |
| 37 | ≤ 6 | F | 1 |  | - | - |  | NP | NP |
| 38 | ≥ 18 | F | 2 |  | - | - |  | NP | NP |
| 39 | ≥ 18 | M | 2 |  | + | + |  | NP | NP |
| 40 | ≤ 6 | M | 2 |  | - | - |  | NP | NP |
| 41 | 7-17 | M | 2 |  | + | + |  | NP | NP |
| 42 | 7-17 | F | 6 |  | - | - |  | NP | NP |
| 43 | ≤ 6 | F | 22 |  | - | - |  | NP | NP |
| 44 | ≤ 6 | F | 2 |  | - | - |  | NP | NP |
| 45 | 7-17 | F | 1 |  | - | - |  | NP | NP |
| 46 | 7-17 | M | 3 |  | - | - |  | NP | NP |
| 47 | 7-17 | M | 3 |  | + | + |  | NP | NP |
| 48 | ≤ 6 | M | 0 |  | - | - |  | NP | NP |
| 49 | ≤ 6 | M | 2 |  | - | - |  | NP | NP |
| 50 | ≤ 6 | M | 3 |  | - | - |  | NP | NP |
| 51 | ≥ 18 | M | 21 |  | - | - |  | NP | NP |
| 52 | ≤ 6 | M | 2 |  | - | - |  | NP | NP |
| 53 | ≤ 6 | F | 2 |  | - | - |  | NP | NP |
| 54 | ≥ 18 | M | 1 |  | - | - |  | NP | NP |
| 55 | ≤ 6 | M | 1 |  | - | - |  | NP | NP |
| 56 | 7-17 | M | 2 |  | - | - |  | NP | NP |
| 57 | 7-17 | F | 1 |  | - | - |  | NP | NP |
| 58 | 7-17 | M | 7 |  | - | - |  | NP | NP |
| 59 | ≥ 18 | M | 3 |  | + | + |  | NP | NP |
| 60 | ≥ 18 | M | 1 |  | - | - |  | NP | NP |
| 61 | ≥ 18 | F | 1 |  | - | - |  | NP | NP |
| 62 | ≤ 6 | F | 1 |  | - | - |  | NP | NP |
| 63 | 7-17 | M | 3 |  | - | - |  | NP | NP |
| 64 | 7-17 | M | 2 |  | - | - |  | NP | NP |
| 65 | 7-17 | M | 1 |  | - | - |  | NP | NP |
| 66 | ≤ 6 | M | 2 |  | - | - |  | NP | NP |
| 67 | 7-17 | M | 3 |  | - | - |  | NP | NP |
| 68 | ≤ 6 | F | 0 |  | - | - |  | NP | NP |
| 69 | 7-17 | M | 1 |  | - | - |  | NP | NP |
| 70 | ≥ 18 | F | 1 |  | + | + |  | NP | NP |
| 71 | ≤ 6 | F | 2 |  | - | - |  | NP | NP |
| 72 | ≥ 18 | M | 0 |  | - | - |  | NP | NP |
| 73 | ≥ 18 | F | 3 |  | - | - |  | NP | NP |
| 74 | ≥ 18 | F | 1 |  | - | - |  | NP | NP |
| 75 | ≥ 18 | F | 5 |  | - | - |  | NP | NP |
| 76 | ≥ 18 | F | 7 |  | - | - |  | NP | NP |
| 77 | ≥ 18 | M | 19 |  | - | - |  | NP | NP |
| 78 | ≥ 18 | F | 0 |  | - | - |  | NP | NP |
| 79 | ≥ 18 | M | 1 |  | - | - |  | NP | NP |
| 80 | ≥ 18 | F | 0 |  | - | - |  | NP | NP |
| 81 | ≥ 18 | M | 8 |  | + | - |  | *C. jejuni* | NP |
| 82 | ≥ 18 | M | 2 |  | - | - |  | NP | NP |
| 83 | ≥ 18 | F | 3 |  | - | - |  | NP | NP |
| 84 | ≥ 18 | M | 4 |  | + | + |  | NP | NP |
| 85 | ≥ 18 | M | 2 |  | + | - |  | - | *C. jejuni* |
| 86 | ≥ 18 | M | 1 |  | - | - |  | NP | NP |
| 87 | ≥ 18 | F | 5 |  | + | + |  | NP | NP |
| 88 | ≥ 18 | M | 7 |  | - | - |  | NP | NP |
| 89 | ≥ 18 | F | 2 |  | + | + |  | NP | NP |
| 90 | ≥ 18 | F | 2 |  | + | + |  | NP | NP |
| 91 | ≥ 18 | F | 2 |  | - | - |  | NP | NP |
| 92 | ≥ 18 | M | 3 |  | - | - |  | NP | NP |
| 93 | ≥ 18 | F | 2 |  | - | - |  | NP | NP |
| 94 | ≥ 18 | M | 3 |  | - | - |  | NP | NP |
| 95 | ≥ 18 | F | 2 |  | - | - |  | NP | NP |
| 96 | ≥ 18 | F | 5 |  | - | - |  | NP | NP |
| 97 | ≥ 18 | F | 3 |  | - | - |  | NP | NP |
| 98 | ≥ 18 | F | 1 |  | + | + |  | NP | NP |
| 99 | ≥ 18 | F | 6 |  | - | - |  | NP | NP |
| 100 | ≥ 18 | F | 0 |  | - | - |  | NP | NP |
| 101 | ≥ 18 | F | 1 |  | - | - |  | NP | NP |
| 102 | ≥ 18 | F | 2 |  | - | - |  | NP | NP |
| 103 | ≥ 18 | M | 10 |  | - | - |  | NP | NP |
| 104 | ≥ 18 | M | 1 |  | - | - |  | NP | NP |
| 105 | ≥ 18 | F | 2 |  | - | - |  | NP | NP |
| 106 | ≥ 18 | M | 0 |  | - | - |  | NP | NP |
| 107 | ≥ 18 | M | 0 |  | + | - |  | - | *C. jejuni*/*C. coli* |
| 108 | ≥ 18 | M | 4 |  | - | - |  | NP | NP |
| 109 | 7-17 | M | 3 |  | - | - |  | NP | NP |
| 110 | ≥ 18 | M | 0 |  | - | - |  | NP | NP |
| 111 | ≥ 18 | M | 1 |  | - | - |  | NP | NP |
| 112 | ≥ 18 | M | 4 |  | + | + |  | NP | NP |
| 113 | ≥ 18 | M | 3 |  | - | - |  | NP | NP |
| 114 | ≥ 18 | M | 6 |  | + | - |  | - | - |
| 115 | 7-17 | M | 6 |  | - | - |  | NP | NP |
| 116 | ≥ 18 | F | 0 |  | - | - |  | NP | NP |
| 117 | ≥ 18 | F | 21 |  | - | - |  | NP | NP |
| 118 | ≥ 18 | F | 1 |  | - | - |  | NP | NP |
| 119 | ≥ 18 | M | 2 |  | - | - |  | NP | NP |
| 120 | ≥ 18 | F | 3 |  | + | + |  | NP | NP |
| 121 | ≥ 18 | M | 1 |  | - | - |  | NP | NP |
| 122 | ≥ 18 | F | 1 |  | - | - |  | NP | NP |
| 123 | ≥ 18 | M | 0 |  | - | - |  | NP | NP |
| 124 | ≥ 18 | F | 0 |  | - | - |  | NP | NP |
| 125 | ≥ 18 | M | 2 |  | - | - |  | NP | NP |
| 126 | ≥ 18 | F | 1 |  | - | - |  | NP | NP |
| 127 | ≤ 6 | F | 1 |  | - | - |  | NP | NP |
| 128 | ≤ 6 | M | 1 |  | - | - |  | NP | NP |
| 129 | 7-17 | M | 2 |  | - | - |  | NP | NP |
| 130 | ≤ 6 | M | 1 |  | - | - |  | NP | NP |
| 131 | ≤ 6 | M | 3 |  | - | - |  | NP | NP |
| 132 | ≤ 6 | F | 2 |  | - | - |  | NP | NP |
| 133 | ≥ 18 | M | 3 |  | + | + |  | NP | NP |
| 134 | ≥ 18 | M | 1 |  | - | - |  | NP | NP |
| 135 | ≥ 18 | M | 8 |  | + | - |  | *C. jejuni* | NP |
| 136 | ≥ 18 | F | 1 |  | - | - |  | NP | NP |
| 137 | ≥ 18 | F | 4 |  | - | - |  | NP | NP |
| 138 | ≥ 18 | F | 1 |  | + | - |  | *C. jejuni* | NP |
| 139 | ≥ 18 | F | 1 |  | + | + |  | NP | NP |
| 140 | ≥ 18 | F | 1 |  | - | - |  | NP | NP |
| 141 | ≥ 18 | M | 2 |  | - | - |  | NP | NP |
| 142 | ≥ 18 | F | 1 |  | - | - |  | NP | NP |
| 143 | ≥ 18 | M | 0 |  | - | - |  | NP | NP |
| 144 | ≥ 18 | F | 1 |  | - | - |  | NP | NP |
| 145 | ≥ 18 | F | 1 |  | + | + |  | NP | NP |
| 146 | ≥ 18 | M | 1 |  | - | - |  | NP | NP |
| 147 | ≥ 18 | F | 2 |  | - | - |  | NP | NP |
| 148 | ≥ 18 | F | 0 |  | - | - |  | NP | NP |
| 149 | ≥ 18 | F | 0 |  | - | - |  | NP | NP |
| 150 | ≥ 18 | M | 1 |  | - | - |  | NP | NP |
| 151 | ≥ 18 | F | 1 |  | + | + |  | NP | NP |
| 152 | ≥ 18 | F | 1 |  | - | - |  | NP | NP |
| 153 | ≥ 18 | F | 3 |  | - | - |  | NP | NP |
| 154 | ≥ 18 | F | 2 |  | + | + |  | NP | NP |
| 155 | ≥ 18 | M | 1 |  | + | + |  | NP | NP |
| 156 | ≥ 18 | F | 2 |  | + | - |  | *C. jejuni* | NP |
| 157 | ≥ 18 | F | 1 |  | + | + |  | NP | NP |
| 158 | ≥ 18 | M | 1 |  | - | - |  | NP | NP |
| 159 | ≥ 18 | F | 4 |  | + | + |  | NP | NP |
| 160 | ≥ 18 | M | 1 |  | + | + |  | NP | NP |
| 161 | ≥ 18 | M | 2 |  | + | - |  | - | - |
| 162 | ≥ 18 | M | 1 |  | - | - |  | NP | NP |
| 163 | ≥ 18 | M | 1 |  | - | + |  | - | - |
| 164 | ≥ 18 | M | 1 |  | - | - |  | NP | NP |
| 165 | ≥ 18 | M | 7 |  | + | + |  | NP | NP |
| 166 | ≥ 18 | M | 6 |  | + | + |  | NP | NP |
| 167 | ≥ 18 | M | 1 |  | - | - |  | NP | NP |
| 168 | ≥ 18 | F | 1 |  | - | - |  | NP | NP |
| 169 | ≥ 18 | M | 1 |  | - | - |  | NP | NP |
| 170 | ≥ 18 | F | 1 |  | + | - |  | - | - |
| 171 | ≥ 18 | F | 2 |  | - | - |  | NP | NP |
| 172 | ≥ 18 | M | 1 |  | + | + |  | NP | NP |
| 173 | ≥ 18 | F | 1 |  | + | + |  | NP | NP |
| 174 | ≥ 18 | M | 1 |  | - | - |  | NP | NP |
| 175 | ≥ 18 | M | 2 |  | - | - |  | NP | NP |
| 176 | ≥ 18 | M | 1 |  | + | + |  | NP | NP |
| 177 | ≥ 18 | M | 1 |  | - | - |  | NP | NP |
| 178 | ≥ 18 | F | 1 |  | - | - |  | NP | NP |
| 179 | ≥ 18 | M | 1 |  | - | - |  | NP | NP |
| 180 | ≥ 18 | M | 1 |  | + | + |  | NP | NP |
| 181 | ≥ 18 | F | 3 |  | + | + |  | NP | NP |
| 182 | ≥ 18 | F | 5 |  | + | + |  | NP | NP |
| 183 | ≥ 18 | M | 1 |  | - | - |  | NP | NP |
| 184 | ≥ 18 | F | 1 |  | - | - |  | NP | NP |
| 185 | ≥ 18 | M | 3 |  | - | - |  | NP | NP |
| 186 | ≥ 18 | M | 1 |  | - | - |  | NP | NP |
| 187 | ≥ 18 | M | 1 |  | - | - |  | NP | NP |
| 188 | ≥ 18 | M | 0 |  | - | - |  | NP | NP |
| 189 | ≥ 18 | M | 1 |  | - | - |  | NP | NP |
| 190 | ≥ 18 | M | 0 |  | - | - |  | NP | NP |
| 191 | ≥ 18 | M | 1 |  | - | - |  | NP | NP |
| 192 | ≥ 18 | M | 1 |  | - | - |  | NP | NP |
| 193 | ≥ 18 | F | 3 |  | - | + |  | - | - |
| 194 | ≥ 18 | M | 1 |  | - | - |  | NP | NP |
| 195 | ≥ 18 | M | 0 |  | - | - |  | NP | NP |
| 196 | ≥ 18 | M | 1 |  | - | - |  | NP | NP |
| 197 | ≥ 18 | M | 1 |  | + | + |  | NP | NP |
| 198 | ≥ 18 | M | 1 |  | - | - |  | NP | NP |
| 199 | ≥ 18 | F | 1 |  | - | - |  | NP | NP |
| 200 | ≥ 18 | M | 1 |  | - | - |  | NP | NP |
| 201 | ≥ 18 | M | 0 |  | - | - |  | NP | NP |
| 202 | ≥ 18 | M | 0 |  | - | - |  | NP | NP |
| 203 | ≥ 18 | M | 1 |  | - | - |  | NP | NP |
| 204 | ≥ 18 | F | 3 |  | - | - |  | NP | NP |
| 205 | ≥ 18 | M | 0 |  | - | - |  | NP | NP |
| 206 | ≥ 18 | M | 1 |  | - | - |  | NP | NP |
| 207 | ≥ 18 | M | 1 |  | - | - |  | NP | NP |
| 208 | ≥ 18 | M | 1 |  | - | - |  | NP | NP |
| 209 | ≥ 18 | M | 2 |  | + | + |  | NP | NP |
| 210 | ≥ 18 | M | 0 |  | - | - |  | NP | NP |
| 211 | ≥ 18 | M | 0 |  | - | - |  | NP | NP |
| 212 | ≥ 18 | F | 0 |  | + | + |  | NP | NP |
| 213 | ≥ 18 | F | 0 |  | - | - |  | NP | NP |
| 214 | ≥ 18 | M | 0 |  | - | - |  | NP | NP |
| 215 | ≥ 18 | M | 3 |  | - | - |  | NP | NP |
| 216 | ≥ 18 | F | 4 |  | - | - |  | NP | NP |
| 217 | ≥ 18 | M | 1 |  | + | + |  | NP | NP |
| 218 | ≥ 18 | M | 2 |  | + | + |  | NP | NP |
| 219 | ≥ 18 | M | 2 |  | + | + |  | NP | NP |
| 220 | ≥ 18 | M | 2 |  | + | + |  | NP | NP |
| 221 | ≥ 18 | M | 0 |  | - | - |  | NP | NP |
| 222 | ≥ 18 | M | 1 |  | + | + |  | NP | NP |
| 223 | ≥ 18 | M | 1 |  | + | - |  | - | *C. jejuni* |
| 224 | ≥ 18 | M | 0 |  | - | - |  | NP | NP |
| 225 | ≥ 18 | F | 2 |  | + | - |  | - | - |
| 226 | ≥ 18 | F | 3 |  | + | + |  | NP | NP |
| 227 | ≥ 18 | F | 3 |  | + | + |  | NP | NP |
| 228 | ≥ 18 | M | 1 |  | - | - |  | NP | NP |
| 229 | ≥ 18 | M | 4 |  | - | - |  | NP | NP |
| 230 | ≥ 18 | M | 7 |  | - | - |  | NP | NP |
| 231 | ≥ 18 | F | 2 |  | + | - |  | *C. jejuni* | NP |
| 232 | ≥ 18 | M | 1 |  | + | + |  | NP | NP |
| 233 | ≥ 18 | M | 4 |  | + | + |  | NP | NP |
| 234 | ≥ 18 | M | 1 |  | - | - |  | NP | NP |
| 235 | ≥ 18 | M | 5 |  | + | + |  | NP | NP |
| 236 | ≥ 18 | M | 1 |  | + | - |  | *C. jejuni* | NP |
| 237 | ≥ 18 | M | 5 |  | - | + |  | - | - |
| 238 | ≤ 6 | M | 1 |  | - | - |  | NP | NP |
| 239 | ≤ 6 | F | 4 |  | - | - |  | NP | NP |
| 240 | 7-17 | M | 1 |  | + | + |  | NP | NP |
| 241 | 7-17 | F | 2 |  | - | - |  | NP | NP |
| 242 | ≤ 6 | F | 1 |  | - | - |  | NP | NP |
| 243 | 7-17 | M | 5 |  | - | - |  | NP | NP |
| 244 | ≤ 6 | F | 5 |  | - | - |  | NP | NP |
| 245 | ≤ 6 | F | 5 |  | + | + |  | NP | NP |
| 246 | ≤ 6 | M | 1 |  | - | - |  | NP | NP |
| 247 | ≤ 6 | M | 28 |  | - | - |  | NP | NP |
| 248 | 7-17 | F | 4 |  | - | - |  | NP | NP |
| 249 | ≤ 6 | F | 1 |  | - | - |  | NP | NP |
| 250 | ≤ 6 | F | 1 |  | - | - |  | NP | NP |
| 251 | 7-17 | M | 3 |  | + | + |  | NP | NP |
| 252 | ≤ 6 | F | 1 |  | - | - |  | NP | NP |
| 253 | ≥ 18 | F | 1 |  | - | - |  | NP | NP |
| 254 | ≤ 6 | M | 3 |  | - | - |  | NP | NP |
| 255 | ≤ 6 | F | 4 |  | - | - |  | NP | NP |
| 256 | ≤ 6 | M | 1 |  | - | - |  | NP | NP |
| 257 | ≤ 6 | M | 1 |  | - | - |  | NP | NP |
| 258 | ≤ 6 | F | 1 |  | - | - |  | NP | NP |
| 259 | ≤ 6 | F | 0 |  | - | - |  | NP | NP |
| 260 | 7-17 | M | 1 |  | - | - |  | NP | NP |
| 261 | ≤ 6 | F | 2 |  | - | - |  | NP | NP |
| 262 | 7-17 | M | 3 |  | + | + |  | NP | NP |
| 263 | ≤ 6 | F | 1 |  | - | - |  | NP | NP |
| 264 | ≤ 6 | F | 1 |  | - | - |  | NP | NP |
| 265 | 7-17 | F | 1 |  | + | + |  | NP | NP |
| 266 | ≤ 6 | M | 3 |  | - | - |  | NP | NP |
| 267 | ≤ 6 | M | 2 |  | - | - |  | NP | NP |
| 268 | ≤ 6 | F | 3 |  | - | - |  | NP | NP |
| 269 | ≤ 6 | M | 4 |  | - | - |  | NP | NP |
| 270 | ≤ 6 | M | 1 |  | - | - |  | NP | NP |
| 271 | ≤ 6 | F | 4 |  | - | - |  | NP | NP |
| 272 | ≤ 6 | F | 5 |  | - | - |  | NP | NP |
| 273 | ≤ 6 | M | 3 |  | - | - |  | NP | NP |
| 274 | 7-17 | M | 1 |  | + | + |  | NP | NP |
| 275 | ≤ 6 | M | 7 |  | - | - |  | NP | NP |
| 276 | ≤ 6 | M | 2 |  | - | - |  | NP | NP |
| 277 | 7-17 | F | 10 |  | - | - |  | NP | NP |
| 278 | ≤ 6 | F | 4 |  | - | - |  | NP | NP |
| 279 | 7-17 | M | 2 |  | + | - |  | - | *C. jejuni* |
| 280 | 7-17 | M | 1 |  | + | + |  | NP | NP |
| 281 | ≤ 6 | F | 5 |  | - | - |  | NP | NP |
| 282 | ≤ 6 | F | 0 |  | - | - |  | NP | NP |
| 283 | ≤ 6 | M | 0 |  | - | - |  | NP | NP |
| 284 | ≤ 6 | M | 1 |  | - | - |  | NP | NP |
| 285 | ≤ 6 | M | 0 |  | - | - |  | NP | NP |
| 286 | ≤ 6 | M | 1 |  | - | - |  | NP | NP |
| 287 | ≤ 6 | M | 1 |  | - | - |  | NP | NP |
| 288 | ≤ 6 | F | 0 |  | - | - |  | NP | NP |
| 289 | ≤ 6 | F | 3 |  | - | - |  | NP | NP |
| 290 | ≤ 6 | M | 1 |  | - | - |  | NP | NP |
| 291 | ≤ 6 | M | 1 |  | - | - |  | NP | NP |
| 292 | ≤ 6 | M | 0 |  | - | - |  | NP | NP |
| 293 | ≤ 6 | M | 1 |  | - | - |  | NP | NP |
| 294 | ≤ 6 | M | 3 |  | - | - |  | NP | NP |
| 295 | ≤ 6 | M | 1 |  | - | - |  | NP | NP |
| 296 | ≤ 6 | M | 3 |  | - | - |  | NP | NP |
| 297 | ≤ 6 | M | 1 |  | - | - |  | NP | NP |
| 298 | ≤ 6 | F | 1 |  | - | - |  | NP | NP |
| 299 | ≤ 6 | M | 7 |  | - | - |  | NP | NP |
| 300 | ≤ 6 | M | 1 |  | - | - |  | NP | NP |
| 301 | ≤ 6 | M | 2 |  | - | - |  | NP | NP |
| 302 | ≤ 6 | M | 2 |  | + | + |  | NP | NP |
| 303 | ≤ 6 | M | 2 |  | - | - |  | NP | NP |
| 304 | ≤ 6 | F | 1 |  | - | - |  | NP | NP |
| 305 | ≤ 6 | F | 3 |  | - | - |  | NP | NP |
| 306 | ≤ 6 | F | 1 |  | - | - |  | NP | NP |
| 307 | ≤ 6 | M | 6 |  | - | - |  | NP | NP |
| 308 | ≤ 6 | M | 1 |  | - | - |  | NP | NP |
| 309 | ≤ 6 | M | 4 |  | + | + |  | NP | NP |
| 310 | ≤ 6 | M | 0 |  | - | - |  | NP | NP |
| 311 | ≤ 6 | F | 0 |  | - | - |  | NP | NP |
| 312 | ≤ 6 | F | 0 |  | - | - |  | NP | NP |
| 313 | ≤ 6 | M | 4 |  | - | - |  | NP | NP |
| 314 | ≤ 6 | M | 3 |  | - | - |  | NP | NP |
| 315 | ≤ 6 | F | 1 |  | - | - |  | NP | NP |
| 316 | ≤ 6 | M | 0 |  | - | - |  | NP | NP |
| 317 | 7-17 | M | 4 |  | + | + |  | NP | NP |
| 318 | ≤ 6 | M | 3 |  | - | - |  | NP | NP |
| 319 | 7-17 | M | 1 |  | + | + |  | NP | NP |
| 320 | 7-17 | M | 1 |  | + | + |  | NP | NP |
| 321 | ≤ 6 | M | 2 |  | - | - |  | NP | NP |
| 322 | ≤ 6 | M | 0 |  | - | - |  | NP | NP |
| 323 | ≤ 6 | M | 2 |  | - | - |  | NP | NP |
| 324 | ≤ 6 | M | 0 |  | - | - |  | NP | NP |
| 325 | ≤ 6 | M | 5 |  | - | - |  | NP | NP |
| 326 | 7-17 | M | 2 |  | + | + |  | NP | NP |
| 327 | 7-17 | M | 0 |  | + | + |  | NP | NP |
| 328 | ≤ 6 | M | 0 |  | - | - |  | NP | NP |
| 329 | ≥ 18 | M | 1 |  | + | + |  | NP | NP |
| 330 | ≤ 6 | F | 0 |  | - | - |  | NP | NP |
| 331 | ≥ 18 | M | 3 |  | - | - |  | NP | NP |
| 332 | ≥ 18 | M | 4 |  | - | - |  | NP | NP |
| 333 | ≥ 18 | M | 4 |  | - | - |  | NP | NP |
| 334 | ≥ 18 | F | 1 |  | - | - |  | NP | NP |
| 335 | ≥ 18 | F | 3 |  | - | - |  | NP | NP |
| 336 | ≥ 18 | M | 1 |  | + | + |  | NP | NP |
| 337 | ≥ 18 | M | 4 |  | - | - |  | NP | NP |
| 338 | ≥ 18 | M | 1 |  | + | + |  | NP | NP |
| 339 | ≥ 18 | F | 5 |  | - | - |  | NP | NP |
| 340 | ≥ 18 | M | 3 |  | + | + |  | NP | NP |
| 341 | ≥ 18 | F | 3 |  | + | - |  | - | *C. jejuni* |
| 342 | ≤ 6 | F | 2 |  | - | - |  | NP | NP |
| 343 | ≥ 18 | M | 1 |  | + | + |  | NP | NP |
| 344 | ≥ 18 | M | 4 |  | + | - |  | - | - |
| 345 | ≥ 18 | F | 1 |  | + | + |  | NP | NP |
| 346 | ≥ 18 | F | 3 |  | - | - |  | NP | NP |
| 347 | ≥ 18 | M | 2 |  | - | - |  | NP | NP |
| 348 | ≥ 18 | F | 3 |  | + | + |  | NP | NP |
| 349 | 7-17 | M | 2 |  | + | + |  | NP | NP |
| 350 | ≥ 18 | F | 1 |  | - | - |  | NP | NP |
| 351 | ≥ 18 | M | 2 |  | + | + |  | NP | NP |
| 352 | ≥ 18 | M | 1 |  | - | - |  | NP | NP |
| 353 | 7-17 | M | 0 |  | + | + |  | NP | NP |
| 354 | ≥ 18 | M | 1 |  | + | + |  | NP | NP |
| 355 | ≥ 18 | M | 3 |  | + | + |  | NP | NP |
| 356 | ≥ 18 | F | 3 |  | + | - |  | - | - |
| 357 | ≥ 18 | M | 1 |  | - | - |  | NP | NP |
| 358 | ≥ 18 | M | 0 |  | + | + |  | NP | NP |
| 359 | 7-17 | M | 1 |  | - | - |  | NP | NP |
| 360 | ≥ 18 | M | 1 |  | - | - |  | NP | NP |
| 361 | ≥ 18 | M | 0 |  | - | - |  | NP | NP |
| 362 | ≤ 6 | F | 1 |  | - | - |  | NP | NP |
| 363 | ≥ 18 | M | 2 |  | - | - |  | NP | NP |
| 364 | ≥ 18 | M | 1 |  | - | - |  | NP | NP |
| 365 | ≥ 18 | F | 2 |  | + | + |  | NP | NP |
| 366 | ≥ 18 | F | 2 |  | - | - |  | NP | NP |
| 367 | ≥ 18 | M | 1 |  | - | - |  | NP | NP |
| 368 | ≥ 18 | F | 0 |  | - | - |  | NP | NP |
| 369 | ≥ 18 | M | 1 |  | - | - |  | NP | NP |
| 370 | ≥ 18 | F | 9 |  | - | - |  | NP | NP |
| 371 | ≥ 18 | F | 1 |  | - | - |  | NP | NP |
| 372 | ≥ 18 | F | 0 |  | - | - |  | NP | NP |
| 373 | 7-17 | F | 1 |  | + | + |  | NP | NP |
| 374 | ≥ 18 | F | 0 |  | - | - |  | NP | NP |
| 375 | ≥ 18 | M | 2 |  | + | + |  | NP | NP |
| 376 | ≥ 18 | F | 1 |  | + | + |  | NP | NP |
| 377 | ≥ 18 | M | 2 |  | - | - |  | NP | NP |
| 378 | ≥ 18 | F | 1 |  | - | - |  | NP | NP |
| 379 | 7-17 | M | 1 |  | - | - |  | NP | NP |
| 380 | ≥ 18 | M | 1 |  | - | - |  | NP | NP |
| 381 | ≥ 18 | M | 1 |  | - | - |  | NP | NP |
| 382 | ≥ 18 | M | 1 |  | + | + |  | NP | NP |
| 383 | ≥ 18 | M | 0 |  | - | - |  | NP | NP |
| 384 | ≥ 18 | M | 0 |  | - | - |  | NP | NP |
| 385 | ≥ 18 | F | 1 |  | - | - |  | NP | NP |
| 386 | ≥ 18 | M | 1 |  | - | - |  | NP | NP |
| 387 | ≥ 18 | F | 0 |  | - | - |  | NP | NP |
| 388 | ≥ 18 | M | 2 |  | + | + |  | NP | NP |
| 389 | ≥ 18 | F | 2 |  | - | - |  | NP | NP |
| 390 | ≥ 18 | M | 0 |  | - | - |  | NP | NP |
| 391 | ≥ 18 | F | 2 |  | - | - |  | NP | NP |
| 392 | ≥ 18 | M | 0 |  | + | - |  | - | - |
| 393 | ≥ 18 | M | 2 |  | - | - |  | NP | NP |
| 394 | ≥ 18 | M | 1 |  | - | - |  | NP | NP |
| 395 | ≥ 18 | M | 2 |  | + | + |  | NP | NP |
| 396 | ≥ 18 | M | 1 |  | + | + |  | NP | NP |
| 397 | ≥ 18 | M | 0 |  | - | - |  | NP | NP |
| 398 | ≥ 18 | M | 1 |  | + | + |  | NP | NP |
| 399 | ≥ 18 | F | 4 |  | + | + |  | NP | NP |
| 400 | ≥ 18 | M | 3 |  | + | - |  | - | - |
| 401 | ≥ 18 | F | 1 |  | + | - |  | - | - |
| 402 | ≥ 18 | M | 0 |  | + | + |  | NP | NP |
| 403 | ≥ 18 | M | 0 |  | + | - |  | - | - |
| 404 | 7-17 | M | 2 |  | + | + |  | NP | NP |
| 405 | ≥ 18 | F | 1 |  | - | - |  | NP | NP |
| 406 | ≥ 18 | M | 1 |  | + | + |  | NP | NP |
| 407 | ≥ 18 | M | 4 |  | - | - |  | NP | NP |
| 408 | ≥ 18 | F | 3 |  | + | + |  | NP | NP |
| 409 | ≥ 18 | M | 1 |  | + | + |  | NP | NP |
| 410 | ≥ 18 | M | 0 |  | - | - |  | NP | NP |
| +, positive; -, negative; NP, not performed | | | | | | | | | |

**Supplementary Figure 1. Melt-Curve Profiles of *Campylobacter* PCR Targets in the FilmArray Gastrointestinal Panel**

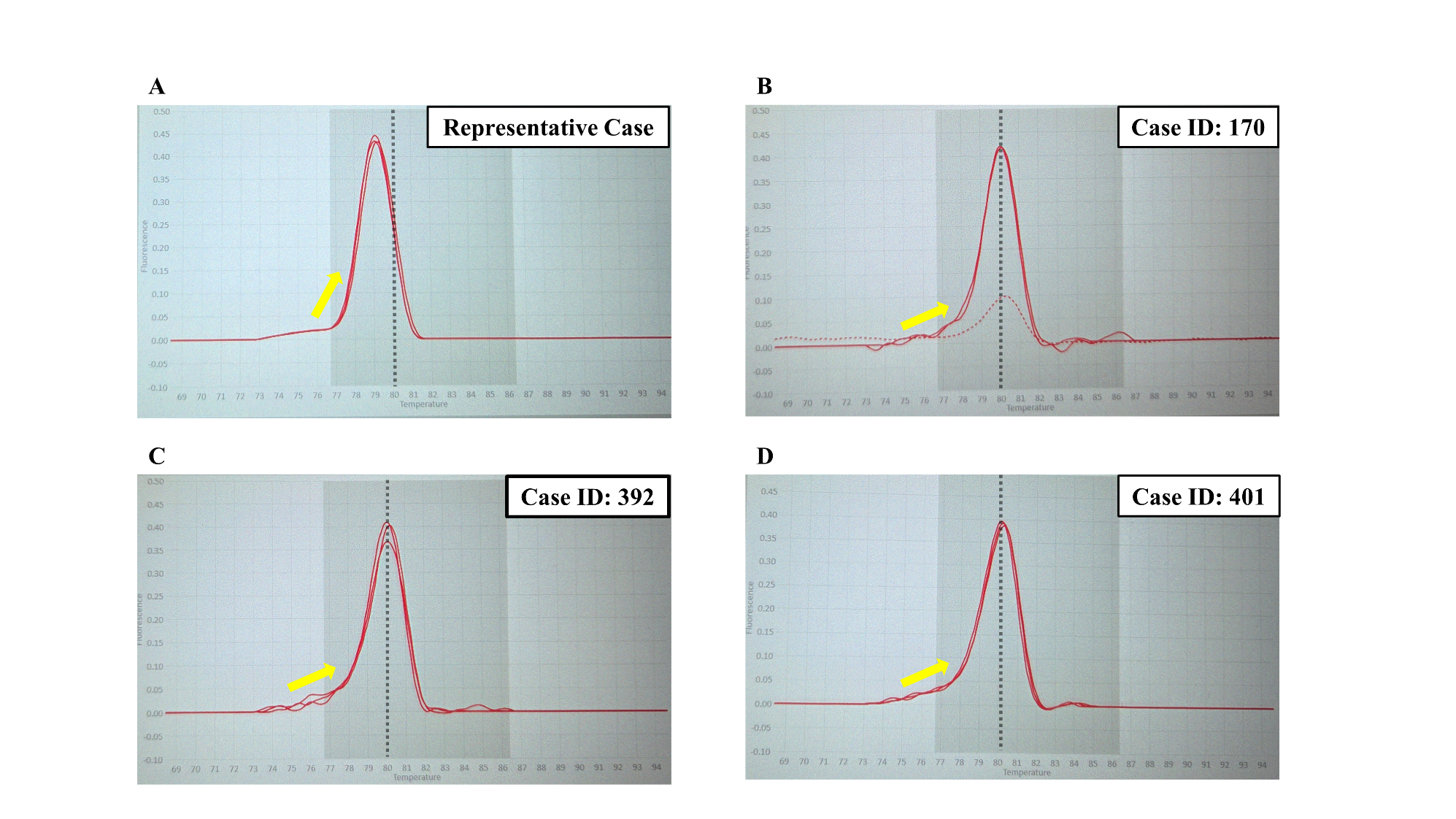

The FilmArray Gastrointestinal Panel detects two Campylobacter-specific targets (Campy 1 and Campy 2) for *Campylobacter jejuni*, *Campylobacter coli*, and *Campylobacter upsaliensis*. A recent study suggested that a characteristic melt-curve pattern of the Campy 2 target may be associated with false-positive results. Panel (A) shows a typical melt-curve pattern observed in a true-positive case. In contrast, panels (B) (Case ID: 170), (C) (Case ID: 392), and (D) (Case ID: 401) exhibit melt-curve patterns suspected to represent false-positive reactions. Relative to the normal pattern in panel (A), which has a melting temperature (Tm) of 79.2 °C, the Tm values in panels (B), (C), and (D) are slightly higher (80.1°C, 80.1°C, and 80.2°C, respectively) and display a prolonged tail toward lower temperatures (left side of the peak), consistent with previously reported findings [20].
